# A Data Driven Change-point Epidemic Model for Assessing the Impact of Large Gathering and Subsequent Movement Control Order on COVID-19 Spread in Malaysia

**DOI:** 10.1101/2020.11.20.20233890

**Authors:** Sarat C. Dass, Wai M. Kwok, Gavin J. Gibson, Balvinder S. Gill, Bala M. Sundram, Sarbhan Singh

## Abstract

The second wave of COVID-19 in Malaysia is largely attributed to a mass gathering held in Sri Petaling between February 27, 2020 and March 1, 2020, which contributed to an exponential rise of COVID-19 cases in the country. Starting March 18, 2020, the Malaysian government introduced four consecutive phases of a Movement Control Order (MCO) to stem the spread of COVID-19. The MCO was implemented through various non-pharmaceutical interventions (NPIs). The reported number of cases reached its peak by the first week of April and then started to reduce, hence proving the effectiveness of the MCO. To gain a quantitative understanding of the effect of MCO on the dynamics of COVID-19, this paper develops a class of mathematical models to capture the disease spread before and after MCO implementation in Malaysia. A heterogeneous variant of the Susceptible-Exposed-Infected-Recovered (SEIR) model is developed with additional compartments for asymptomatic transmission. Further, a change-point is incorporated to model the before and after disease dynamics, and is inferred based on data. Related statistical analyses for inference are developed in a Bayesian framework and are able to provide quantitative assessments of (1) the impact of the Sri Petaling gathering, and (2) the extent of decreasing transmission during the MCO period. The analysis here also quantitatively demonstrates how quickly transmission rates fall under effective NPI implemention within a short time period.

## 1 Introduction

Imported cases from China contributed to the first COVID-19 wave in Malaysia from January 25, 2020 to February 26, 2020 [1]. This first wave had a total of 22 cases out of which 20 were directly linked to foreign travel while the remaining two cases were local transmissions [2, 3]. The second wave of COVID-19 in Malaysia was largely attributed to a mass gathering held in Sri Petaling between February 27, 2020 and March 1, 2020, which contributed to an exponential rise of COVID-19 cases in the country. This gathering involved over 16, 000 participants including a large number of foreigners from countries that later registered COVID-19 cases [4]. On March 18, 2020, the Malaysian government introduced a nationwide lockdown which was the Phase I Movement Control Order (MCO) throughout the country to stem the spread of COVID-19. Phase 1 MCO was enforced for a 2-week period starting from March 18, 2020 to March 31, 2020. During this phase, various non-pharmaceutical interventions (NPIs) were strictly enforced by means of movement restrictions, wearing of face masks, social distancing and hand hygiene practices to reduce disease transmission. The Phase I MCO was then evaluated after 2 weeks based on case trends and model forecasts by the Ministry of Health (MOH) Malaysia. As local transmission persisted, the MCO was extended a total of four times until May 12, 2020. Phase 2, 3 and 4 of the MCO covered periods starting from April 1, 2020 until May 12, 2020 (Phase 2 - April 1, 2020 to April 14, 2020, Phase 3 - April 15th to April 30, 2020, and Phase 4 - May 1, 2020 to May 12, 2020). Subsequently, from May 4, 2020, the MCO was eased into the Conditional MCO (CMCO) until June 9, 2020. However, during CMCO, there were still several identified hot-spots of COVID-19 which were placed under Enhanced MCO (EMCO) with the aforementioned strict movement control restrictions. The implementation of MCO proved to be effective - the reported COVID-19 cases reached its peak around the first week of April and subsequently started to reduce. However, concerns remained whether a rebound in transmission would occur when the MCO was lifted and if compliance to NPIs were not followed strictly at that time. In order to gain a quantitative understanding of the effect of MCO, we developed a class of mathematical models to capture the dynamics of COVID-19 spread before and after the MCO implementation. A variant of the Susceptible-Exposed-Infected-Recovered (SEIR) model is proposed and developed in this paper which incorporates heterogeneity in the transmission dynamics, additional compartments for asymptomatic transmission and a change-point, chosen adaptively based on data, to reflect the shift in spread dynamics after the MCO implementation. The models developed in this paper are able provide a quantitative assessment of the extent of COVID-19 spread during the pre-MCO (large gathering) and MCO periods by means of a measure of infectivity developed from them. This measure is similar to the basic reproduction number, commonly denoted by ℛ_0_, but can be calculated for more complex epidemic models such as the ones proposed here.

Deterministic compartmental models, such as the Susceptible-Infected-Recovered (SIR) or the Susceptible-Exposed-Infectious-Recovered (SEIR) models, provide a good theoretical framework to study infectious disease spread, and have been widely used and reported in the literature. However, more complex versions of these models, and their stochastic counterparts require data-rich inputs to model all aspects of the disease dynamics. Data-rich inputs, if lacking, can be compensated using reliable and informative prior elicitation. Considering the acute nature and scale of the pandemic as well as the urgent need for a multisectorial response, comprehensive data availability of the pandemic was limited in Malaysia. For example, the open source website `outbreak.my` initially reported a transmission network for all cases; however, it was unable to cope with the scale of the pandemic when it intensified. Factoring in this data limitation, we choose to develop models that are deterministic, rather than stochastic, while ensuring that they are able to capture salient transmission dynamics satisfactorily. As mentioned earlier, we enhance the deterministic models by incorporating compartments for asymptomatic transmission and a change-point to reflect the shift in disease dynamics. We also take into account heterogeneity in the disease spread such as varying contacts among susceptibles within the closed population. The starting point of our proposed models are the class of epidemic models with power transmission dynamics which are shown to incorporate heterogeneity (see [5, 6]).

Several studies in the literature [7, 8, 9, 10, 11] have analyzed the effects of NPIs in reducing the number of COVID-19 cases. In [7], the effects of different types of NPIs on COVID-19 cases are modeled using a negative-binomial distribution whose underlying parameters incorporate country information, type of NPI implemented and change-point effects. The associated Bayesian analysis is carried out using Markov Chain Monte Carlo (MCMC) algorithms to arrive at posterior parameter estimates and credible intervals. No epidemic models are considered in this work. A generalization of the SEIR epidemic model is considered in [9] to understand the dynamics of transmission in New York, USA, under various NPI settings. However, the model is complex and requires data-rich inputs for the estimation of all unknown parameters. As a result, the authors derive baseline epidemiological parameters from published literature and not from actual observed cases in New York, and in the end conduct a simulation study based on the assumed parameter values. The study in [8] extends the work of [12] and computes a time-varying basic reproduction number as a way of gauging the effect of NPIs over time. Both these works assume that serial intervals (i.e., the time between onset of symptoms for the infector and the infectee) can be computed for each case, which is another situation requiring data-rich input.

Studies that use compartmental epidemic models as a way of gauging the time-varying effects of NPIs have emerged over the course of the pandemic [13, 14, 15]. Compartmental epidemic models naturally model disease spread via contact rates which directly quantify the extent of NPIs (since, as mentioned earlier, NPIs are designed to reduce person-to-person transmission). Thus, epidemic models provide a natural approach for considering time-varying effects of the MCO period. Further, in this paper, the estimation of SEIR parameters is carried out based on local considerations and local data; they are not obtained from published literature based on studies conducted elsewhere where their local dynamics can be vastly different.

We seek to address one important aspect of Malaysia’s multifaceted response to the COVID-19 pandemic, that is, to inform the health officials at MOH and aid them in their decision-making. Thus, our model was developed under local considerations using local data. Our model and related analyses are able to provide a quantitative assessment of (1) the impact of the Sri Petaling gathering, and (2) the extent of decreasing transmission during the MCO period by incorporating a time-varying contact rate parameter, which is estimated using locally available data. In essence, the proposed models here are being used as a lens to interpret the observed data in terms of when, and to what extent, a reduction in COVID-19 transmission occurred as result of the implementation of MCO.

The remainder of the paper is organized as follows: Section 2 presents the material and methods used in this paper: Section 2.1 gives the various data sources used in this study, Section 2.2 describes the epidemic models that we propose in this paper and Section 2.5 outlines the related Bayesian inference methodology developments. Section 3 gives the results of our analyses, and Section 4 provides general discussion and insights derived from these results. Finally, several conclusions and potential future work are outlined in Section 5.

## 2 Methods

### 2.1 Data Collection

Daily situation reports on COVID-19 cases in Malaysia are published by the National Crisis Preparedness and Response Centre (CPRC) of MOH, as well as other official websites (such as `outbreak.my`). The data on daily COVID-19 cases have been published since 21 January 2020 and is publicly available. The reports consist of confirmed daily and cumulative cases, recovered cases and deaths, as well as cases requiring ICU care and ventilator support. Cases by states are also available for the 13 states and 3 federal territories. In this study, we studied characteristics of the second COVID-19 wave in Malaysia starting from March 1, 2020 (corresponding to the final day of the Sri Petaling gathering). Data used for the current study are confirmed daily cases for Malaysia, and for two states: Selangor and Sarawak. These states were chosen to illustrate the aggressive transmission propagated by the Sri Petaling gathering. Selangor is the state where Sri Petaling is located and from where a majority of the participants originated, whereas Sarawak represents a state which was essentially not affected by this gathering. The time period of study is between March 1, 2020 (end of Sri Petaling gathering) and April 28, 2020, covering the period immediately after the Sri Petaling gathering and the first three phases of the MCO. Our study duration is further divided into two periods. The first period ranges from March 1, 2020 until March 18, 2020, which covers the subsequent 17 days after the gathering. The second period is taken from March 18, 2020 until April 28, 2020, covering the three successive Phases 1, 2 and 3 of the MCO. Figure 1 gives the trajectories of reported daily COVID-19 cases between March 1, 2020 and April 28, 2020 for Malaysia, and the states of Selangor and Sarawak.

**Figure 1:**
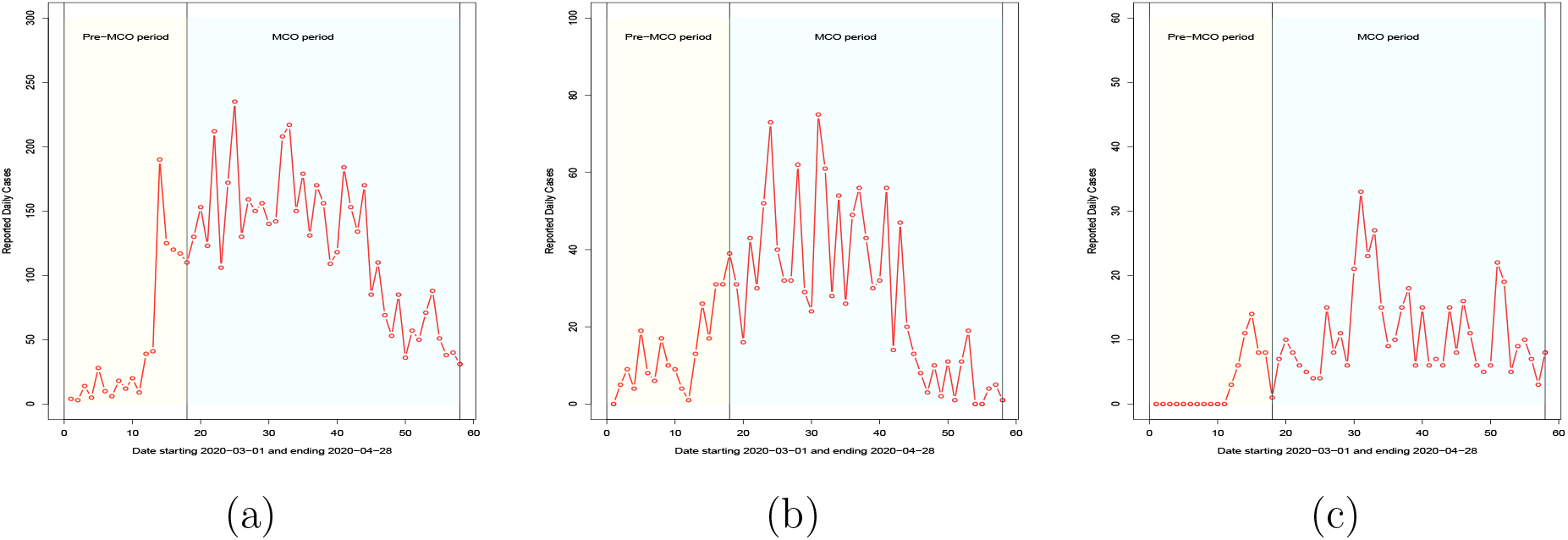
Reported daily cases for (a) Malaysia, (b) Selangor and (c) Sarawak. The time period considered is from March 1, 2020 to April 28, 2020.

All COVID-19 cases reported by MOH were confirmed by real-time reverse transcriptase-polymerase chain reaction (real-time RT-PCR) tests. A positive case was reported when the person in question was found to be positive for SARS-CoV-2 via a real-time RT-PCR test. Upon confirmation, the individual was isolated at COVID-19 designated hospitals and healthcare facilities. Active cases are defined as infected persons who were currently undergoing treatment, and hence, isolated and removed; the individual is assumed to be unable to infect other susceptibles in the population, and hence “removed” from further modelling steps. This study did not consider transmission from positive isolated COVID-19 patients to health personnel as there was no evidence of this type of transmission occurring in the COVID-19 designated hospitals in Malaysia.

### 2.2 The SEIR model

The typical and well-known SEIR compartmental model consists of four compartments (Susceptible, Exposed, Infected and Recovered) representing different stages of evolution of an infectious disease, such as COVID-19, in a population. Susceptible individuals come in contact with one or more infected individuals in the population, and subsequently, become exposed to the virus. The virus then incubates within these individuals for some time. At the end of the incubation period, the exposed person becomes infectious and transmits the disease to other susceptibles in the population who come in close contact to him/her. The infected person is assumed to be infectious for a certain period (called the infectious period) after which the person recovers, dies or becomes immune. The deterministic SEIR model is given by a set of nonlinear ordinary differential equations (ODEs):

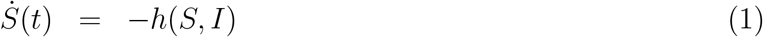

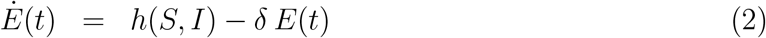

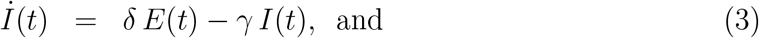

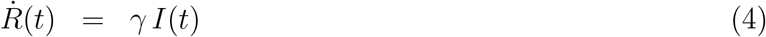

where *S*(*t*), *E*(*t*), *I*(*t*) and *R*(*t*) represents, respectively, the susceptible, exposed, infected and recovered compartments representing the total number of individuals in each compartment at time *t* (here, 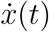 denotes the derivative of *x*(*t*) with respect to time *t* for *x ∈ {S, E, I, R}*), *N* is the total population size, and 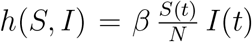 is the rate of new infections (or, the number of new cases in the population). The parameters that govern the trajectory of the SEIR model are *θ ≡* (*β, δ, γ, i*_0_, *e*_0_) which are, respectively, the transmission rate (i.e., number of individuals in the population an infected person comes in contact with and successfully transmits the disease per unit time), the rate of incubation of the disease, the rate of infectiousness, the initial number of infectives and the initial number of exposed individuals. Since 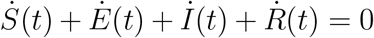, it follows that *S*(*t*) + *E*(*t*) + *I*(*t*) + *R*(*t*) = *N* for all *t*. Reparametrizing *S*(*t*) = *S*(*t*)*/N, E*(*t*) = *E*(*t*)*/N, I*(*t*) = *I*(*t*)*/N* and *R*(*t*) = *R*(*t*)*/N*, the renormalized versions of *S, E, I* and *R* represent the proportion, rather than the total number of individuals, in each compartment. In the renormalized SEIR model, *S*(*t*) + *E*(*t*) + *I*(*t*) + *R*(*t*) = 1 and the rate of new infections become

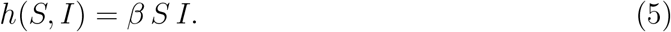

Based on initial values of 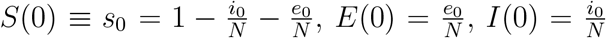, and *R*(0) = 0 at time *T*_0_ = 0, the SEIR ODE system can be solved numerically to yield the values of *S*(*t*), *E*(*t*), *I*(*t*) and *R*(*t*) for every *t ∈* [*T*_0_, *T*_1_] where *T*_1_ denotes the final time-point. In (1)-(4), the incubation period, 1*/δ*, is inversely proportional to the incubation rate *δ*, and similarly, the infectious period 1*/γ* is inversely proportional to the infectious rate *γ*. The correspondence between rates and exponential sojourn times is only approximately so since it is not so straightforward to establish this correspondence with an individual-based stochastic model given the non-linear nature of the process.

Modifications to typical SEIR formulation (1)-(4) are made to adapt it to the local Malaysian context. Here, the last compartment of the SEIR model is not “Recovered” but “Removed”, representing all infectious individuals who are effectively isolated following a positive test result. In Malaysia, such patients are isolated in hospital wards to avoid further contacts with susceptibles. For COVID-19 in particular, the onset of symptoms does not necessarily indicate the start of infectiousness; in fact, the onset of infectiousness may be somewhat earlier. Thus, the infectious period 1*/γ* represents the period of effective infectiousness, that is, the period from the start of infectiousness (asymptomatic or symptomatic) until the individual is isolated and then can no longer infect others. Based on this understanding, 1*/δ* represents the incubation period, which is the period starting from getting infected until the onset of infectiousness. The connection between the SEIR modelling formulation and actual evolution stages of COVID-19 in patients is shown in Figure 2. Recent studies on COVID-19 have clearly reported growing evidence of asymptomatic infections [16, 17, 18] which the current SEIR model does not incorporate. We address the issue of asymptomatic transmission in our subsequent model development in Section 2.3.

**Figure 2:**
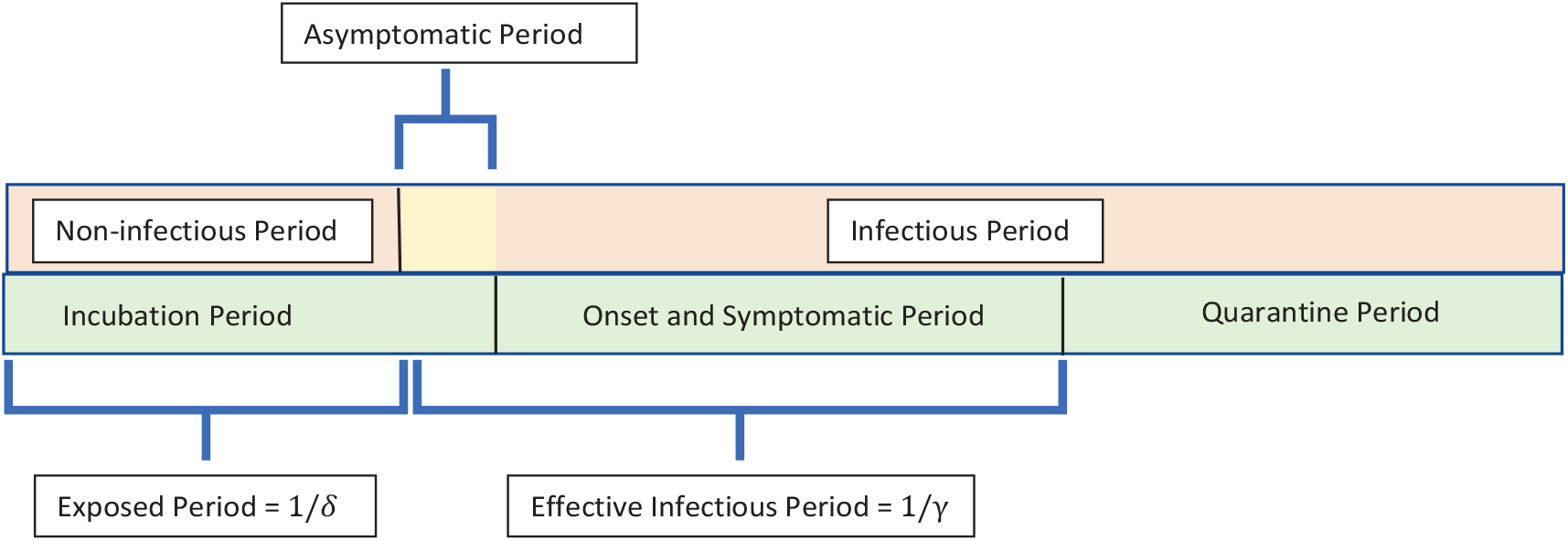
Salient periods in the evolution of COVID-19 and their connection to durations in the SEIR model.

Observed data in Malaysia consists of the total number of confirmed daily cases as reported by `outbreak.my` and CPRC (i.e., the number of cases that were tested positive, and hence, effectively isolated). Hence, only the *R* compartment of the SEIR model can be related to observed data while other compartments of the SEIR model remain unobserved. In the subsequent model development in Section 2.3, the *R* compartment is further split into two: observed and cryptic sub-compartments corresponding to reported and asymptomatic infections; see Section 2.3 for more details. An additional assumption made is that individuals from the *R* compartment do not return to the *S* compartment - at least on the timescales over which the epidemic is observed; see, for example, [19, 20]

In order to provide a quantitative assessment of the impact of the MCO, we modify (1)-(4) to incorporate an instantaneous time-varying transmission rate, *β ≡ β*(*t*) (see [21], for example) which is able to quantify the extent of disease transmission at time *t*. The MCO can be deemed effective if the function *β*(*t*) shows a decay reflecting an increasing effectiveness in reducing transmission among individuals over time due to implementation of the NPIs.

### 2.3 The modified SEIR model

The aforementioned SEIR model does not account for heterogeneity, asymptomatic transmission and change points. To this end, we propose models with power transmission dynamics that incorporate heterogeneity in the disease parameters; see, for example, [5, 6]. Further, the infectious compartment in (3) of the SEIR model is now split into two, *I*_*o*_ and *I*_*c*_, for symptomatic (or, observed) and asymptomatic (or, cryptic) individuals, who, respectively, exhibit and do not (or, mildly) exhibit symptoms but are nevertheless infectious. Correspondingly, the *R* compartment is also split into two, as mentioned earlier, to accomodate quarantined and un-quarantined cases. It is assumed that the proportion of individuals transiting from *E* to *I*_*o*_ is *p*. The remaining exposed individuals (a proportion of 1 *− p*) transition into the *I*_*c*_ compartment and remain undetected throughout their disease experience.

From now on, we consider only renormalized state values which represent proportions, and not actual numbers, of the population. The modified SEIR model with cryptic and observed infectiousness is given by the following system of ODEs:

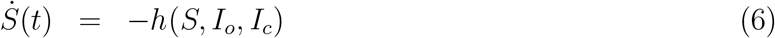

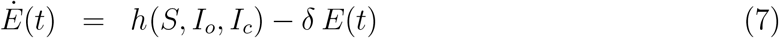

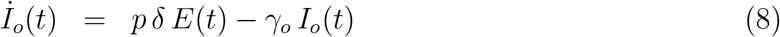

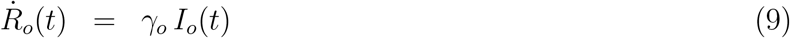

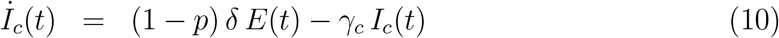

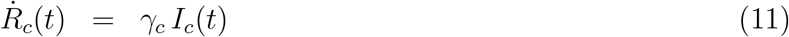

where the rate of new infections is now given by

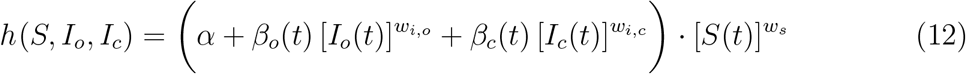

as opposed to *β S*(*t*) *I*(*t*) in (5) for the SEIR model. A key difference of the model formulation in (6) - (12) is the power transmission dynamics used to model heterogeneity in the population; see [5, 6]. In [6], a gamma distribution is elicited on the varying disease transmission parameters within the population which gives rise to the powers 𝒲_*s*_, 𝒲_*i,o*_ and 𝒲_*i,c*_ on *S*(*t*), *I*_*o*_(*t*) and *I*_*c*_(*t*), respectively, with 𝒲_*s*_ *≥* 1, 𝒲_*i,o*_ *≥* 1 and 𝒲_*i,c*_ *≥* 1. The lower bounds on 𝒲_*s*_, 𝒲_*i,o*_ and 𝒲_*i,c*_ recover the original SEIR model dynamics with no heterogeneity. The remaining parameters have the following interpretation: (1) *α* represents a small yet significant force of infection that starts the local infection process but is eventually overwhelmed by it. In our context, *α* represents the initial force of infection arising from, say, foreign infectious individuals attending the large gathering at Sri Petaling starting February 27th. (2) The parameters *γ*_*o*_ and *γ*_*c*_ have the same interpretation as *γ* in (3), that is, they are rates of infectiousness but for the observed and cryptic compartments, respectively. (3) The parameter *δ* is the same as before, namely, it is the rate of incubation associated with the exposed compartment. (4) The parameters *β*_*o*_ and *β*_*c*_ are transmission rates for the observed and cryptic compartments, respectively, in the modified SEIR model with *β*_*c*_ = *µ β*_0_ and *µ ∈* [0, 1]. In other words, we assume that the transmission rate for asymptomatic individuals is smaller than that of symptomatic individuals; this is a plausible assumption to make as asymptomatic individuals generally possess a lower viral load which leads to lower chances of a successful transmission. On the other hand, a longer asymptomatic infectious period may compensate for the lower transmission rates for an asymptomatic individual, and this possibility is captured by the model via the parameter *γ*_*c*_.

A change-point is incorporated into the modified SEIR model to capture the shift in disease dynamics before and after the start of the MCO. For this, an unknown threshold, *T*^*\**^, is chosen so that observed daily cases fall either to the left or right of *T*^*\**^. We denote the observation window to the left of *T*^*\**^ by 𝒲_*L*_ which consists of dates from March 1st up to and including *T*^*\**^. The window to the right of *T*^*\**^ is denoted by 𝒲_*R*_ which consists of dates after *T*^*\**^ up to and including April 28th, 2020. The change-point date *T*^*\**^ is inferred from data; it is not taken as March 18th, 2020, the date of the start of MCO. Choosing *T*^*\**^ in this data-driven way is justified as the impact of MCO on observed cases may not be realized immediately. The modified SEIR model with change-point *T*^*\**^ is governed by the ODE system (6)-(12) for all time points *t ∈ 𝒲*_*L*_. For *t ∈ 𝒲*_*R*_, the same ODE system (6)-(12) is considered but now with a different set of parameter values in 𝒲_*R*_ compared to 𝒲_*L*_, and with a new *β*_*o*_ expressed as a function of *t*,

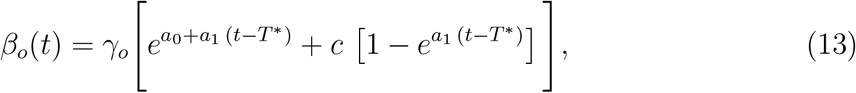

to model possible changes in disease transmission over time. The functional form of *β*_*o*_(*t*) in (13) has an initial value of 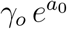 at *t* = *T*^*\**^ after which it decreases (provided *a*_1_ *<* 0) to the asymptotic value of *cγ*_*o*_ as *t → ∞*. Thus, *cγ*_*o*_ represents the residual disease transmission that may be present even during the MCO period, for example, due to close contact between family members in the same household. The general functional form of *β*_*o*_(*t*) subsumes the constant disease transmission rate model as a special case by taking *a*_1_ = *c* = 0 in (13). The constant rate submodel has the advantage of not explicitly assuming any functional form for the change in disease transmission over time. On the other hand, it can only ascertain if there is an overall change (increase or decrease) in transmission after the change point *T*^*\**^. Similar to the relationship *β*_*c*_ = *µβ*_*o*_ in 𝒲_*L*_, we assume *β*_*c*_(*t*) = *µ β*_*o*_(*t*) for the window 𝒲_*R*_ based on a different *µ* value. In Section 3, the submodel is used first followed by the full model to fit to observed data.

In the model formulation of (6)-(11), only the *R*_*o*_ compartment is modelled directly using a likelihood function based on daily observed cases. The other compartments of the modified SEIR model remain latent and do not have any direct observation processes for modelling based on likelihoods; see Section 2.5 for further details.

### 2.4 Quantitative assessment of disease spread

The basic reproduction number, ℛ_0_, is defined as the number of secondary infections caused by one primary individual during his/her infectious period. It is the most important quantitative indicator reported to assess whether the disease is in control or not. It is well-known that the threshold value of 1 for ℛ_0_ distinguishes between the situations where a major epidemic occurs versus the disease dying out eventually in the population. In fact, several studies in the literature report a gradual decrease in ℛ_0_ after lockdown [22, 23, 24]. For the SEIR model in (1) - (4), ℛ_0_ is given by the well-known formula ℛ_0_ = *β/γ*. Time-varying measures of secondary infections per primary infected, ℛ_*t*_, are also available in the literature. However, for the modified SEIR model presented in Section 2.3, ℛ_0_ and ℛ_*t*_ cannot be computed. Therefore, we resort to an alternative quantitative assessement of disease spread - the total number of infections (i.e., generational) caused by the introduction of one additional infectious individual into the infection process at time point *t*. This procedure is illustrated using the *I*_*o*_ compartment. Based on (6)-(12), new values for the ODE system are calculated from time point *t* onwards with current state values serving as initializations of the ODE system for all except one compartment: For the *I*_*o*_-compartment, the current value *I*_*o*_(*t*) is replaced by *I*_*o*_(*t*) + 1*/N* as the initial value. The new rate of incidence is given by 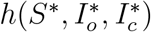 over the infectious period of the individual when the ODE system is propagated using (6)-(12) in [*t, t* +1*/γ*_*o*_]. The increase in the rate of incidence by the introduction of this individual in the *I*_*o*_ compartment is given by

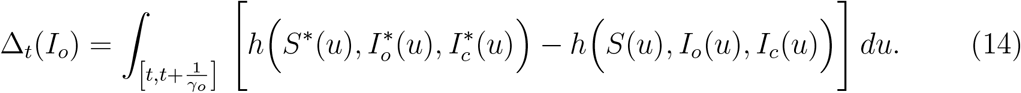

Similarly, the increase in the rate of incidence by the introduction of one infectious individual into the *I*_*c*_ compartment at time point *t* can be calculated. This is denoted by Δ_*t*_(*I*_*c*_). The final increase in the incidence is the mixture

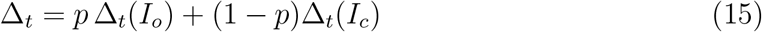

where *p* is the proportion of exposed individuals who enter the *I*_*o*_ compartment in (8).

### 2.5 Bayesian Inference

Model fitting and inference is carried out in a Bayesian framework.

### 2.5.1 The Likelihood

The likelihood relating components of the *R*_*o*_-compartment to the total number of daily cases, *D*_*t*_, *t ∈* 𝒲_*L*_ *∪* 𝒲_*R*_, is taken to be the negative binomial probability function, that is,

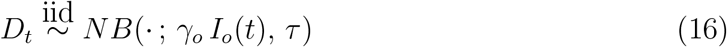

Where

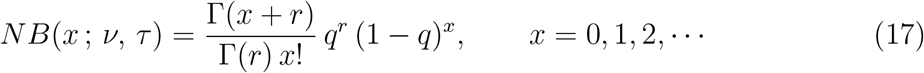

has mean *ν ≡ q r/*(1 *− q*) and variance *qr/*(1 *− q*)^2^ *≡ ν* + *τ ν*^2^; in (17), Γ(*u*) is the Gamma function 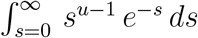 *ds* evaluated for *u >* 0. Thus, *r* need not be integer-valued in (17). The parameter *τ* measures overdispersion with respect to the Poisson likelihood recovered when *τ* = 1; see, for example, [25, 26]. The observed data on daily cases in Malaysia exhibited significant overdispersion in the order of the mean. As a result, the Poisson likelihood did not fit well and we resorted to the negative binomial likelihood instead. More discussion on this aspect is presented later.

### 2.5.2 Assignment of Priors

Let 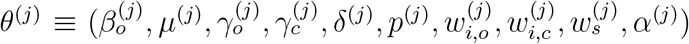 for *j* = *{L, R}* be the parameters of the ODE system (6)-(12) for *t ∈ 𝒲*_*L*_ and *t ∈ 𝒲*_*R*_, respectively. The collection of all parameters is denoted by Θ = *θ*^(*L*)^ *∪ θ*^(*R*)^ *∪* (*τ, T*^*\**^) where *τ* is the overdispersion parameter of the negative binomial likelihood and *T*^*\**^ is the change-point. The uncertainty in *θ* is elicited via prior distributions in the Bayesian inferential framework. In what follows, we describe the priors used on the generic parameter *θ* after which the discussion on prior elicitation can be extended to *θ*^(*L*)^ and *θ*^(*R*)^ in a straightforward manner. It is important to note that the priors on *θ* depend on hyperparameters. Here, we only elicit the forms of the priors; the discussion on the exact choice of the corresponding hyperparameters is relegated to the next few paragraphs. Independent uniform priors 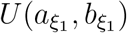 are chosen for each *ξ*_1_ *∈ {µ, p, w*_*i,o*_, 𝒲_*i,c*_, 𝒲_*s*_, *α}* with corresponding hyperparameters 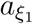 and 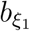. The prior on (*γ*_*o*_, *δ*) is chosen independently in the following way: For 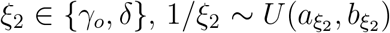. The reciprocal transformation is used for prior elicitation since these parameters have a rate interpretation, and hence, their reciprocals represent the corresponding duration (either incubation or infectious periods) with benchmark values reported in the literature [16, 17, 18, 27]. The cryptic infectious period 1*/γ*_*c*_ is generally longer than the observed infectious period 1*/γ*_*o*_ (since the former remains undetected). Hence, it follows that 1*/γ*_*o*_ *<* 1*/γ*_*c*_. This restriction can be incorporated into the prior elicitation for 1*/γ*_*o*_ and 1*/γ*_*c*_ by first generating 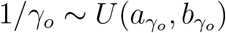 as above and then setting 1*/γ*_*c*_ = 1*/γ*_*o*_ + *ξ*_3_ where 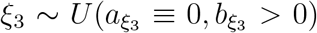 *>* 0). The prior on the change point *T*^*\**^ is taken to be uniform on dates from March 18th 2020 to March 31st 2020 both inclusive. Since we define *T*^*\**^ as the number of days after March 1st 2020, *T*^*\**^ *∼ U* (17, 30). The prior on *τ* is taken to be *U* (*a*_*τ*_, *b*_*τ*_) for the entire observation window 𝒲_*L*_ *∪* 𝒲_*R*_.

The prior on *β*_*o*_ is elicited indirectly via a reparametrization: For *𝒲*_*L*_, we take 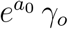 and consider a uniform prior on *a*_0_, independent of *γ*_*o*_. Our prior elicitation adopts this reparametrization for the following reason: For 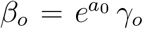, the basic reproduction number 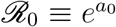 is the more fundamental quantity for simpler models, such as the SIR and SEIR models, compared to *β*_*o*_. Thus, *R*_0_ for these models can be benchmarked based on similar flu-like epidemics in the past which, in turn, provide a suggested range of values for the prior elicitation of *a*_0_ in 𝒲_*L*_; see [28]. Note that *β*_*o*_ which represents the contact rate of the target population cannot be determined by direct observation. Hence, putting a prior directly on *β*_*o*_ is difficult. Similarly, since 1*/γ*_*o*_ is the observed infectious period, benchmark values can be obtained from the literature for its prior elicitation. To summarize, we put direct priors on parameters that have an epidemiological interpretation and which can be benchmarked from reported literature for eliciting the appropriate prior.

For 𝒲_*R*_, *β*_*o*_(*t*) has the form in (13) for coefficients *a*_0_, *a*_1_ and *c*. For *a*_0_, we consider two cases: When using the submodel to determine whether an overall reduction in transmission occurs or not, we take 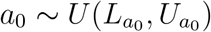 with 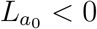 and 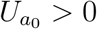. If the full model of (13) is considered, *a*_0_ is chosen deterministically to ensure continuity of the infection process before and after the change-point. More specifically, *a*_0_ in 𝒲_*R*_ is chosen so that the rate of incidence *h*(*S*(*t*), *I*_*o*_(*t*), *I*_*c*_(*t*)) in 𝒲_*L*_ and 𝒲_*R*_ coincide at *t* = *T*^*\**^. For the full model, *a*_1_ is given a uniform prior with support on 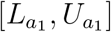. We take 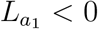 and 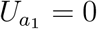 if such a reduction in disease transmission is established by the submodel.

Prior elicitation on parameters in 𝒲_*R*_ is based on convenience and ease of interpretation. It is easier to elicit priors on the components of *β*_*o*_(*t*) in (13) compared to the entire function itself. Further, the component parameters of *β*_*o*_(*t*) satisfy restrictions that are easy to understand; for example, 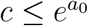 since *c* and 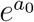 are the lower and upper bounds of the decay curve, respectively. Hence, a reasonable prior to put on *c* is 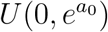. The prior elicitation on remaining unknown parameters are explained in detail in the Appendix.

#### 2.5.3 Computational algorithm

Based on the negative binomial likelihood and prior elicitation in Sections 2.5.1 and 2.5.2, respectively, the posterior of Θ can be derived using Bayes theorem as

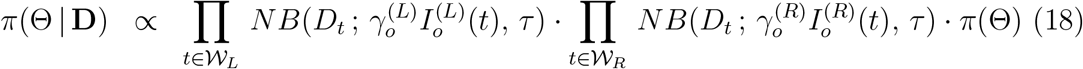

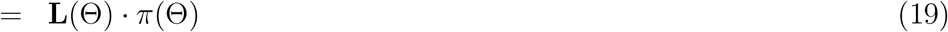

where **D** = *{D*_*t*_ : *t ∈* 𝒲_*L*_ *∪* 𝒲_*R*_*}* is the collection of observed daily cases from March 1st until April 28, 2020, **L**(Θ) is the entire likelihood for **D** and *π*(Θ) is the prior on Θ as described in Section 2.5.2. Bayesian inference is carried out using Monte Carlo importance sampling. A total of *M* samples, Θ_*i*_ for *i* = 1, 2, …, *M*, are generated from the prior specification *π*(Θ). The likelihood, **L**(Θ_*i*_), is computed for each Θ_*i*_ and normalized to obtain weights

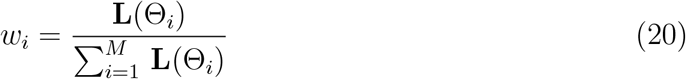

for *i* = 1, 2, *…, M*. To compute **L**(Θ_*i*_), one requires to numerically evaluate the solution of the ODE system (6)-(12). This is achieved using the `deSolve` package in R. The Bayesian computational algorithm described here is developed using R and the Rstudio® user interface. Through this importance sampling step, an approximation to the Maximum-a-Posteriori (MAP) estimator of Θ is given by

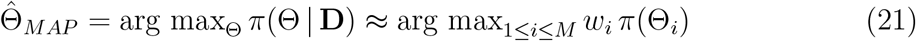

with the approximation becoming more accurate as *M → ∞*. Resampling Θ_*i*_s with weights 𝒲_*i*_ in (20) gives a ensemble of size *M*, 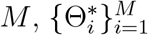, from the target posterior *π*(Θ | **D**) which can be used to provide a quantification of uncertainty around 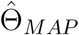.

## 3 Results

The study period is from March 1, 2020 until April 28, 2020 with *T*_0_ = 0 and *T*_1_ = 59. The impacts of the Sri Petaling gathering and MCO implementation are analyzed here using the proposed model described in Section 2.3. Reported daily cases at the national level as well as for Selangor and Sarawak are used for model fitting and parameter estimation. Note that all states in Malaysia implemented MCO Phases 1-3 using the same guidelines and protocols. Thus, one can gauge the impact of the Sri Petaling gathering on COVID-19 spread in Malaysia based on a comparison between states and the national experience. Here, Selangor and Sarawak are chosen as two such representative states with high and low population densities, respectively.

First, we investigate if the MCO implementation had an overall effect of reducing COVID-19 transmission rates. For this purpose, the constant rate submodel of (13) is used and the prior on *a*_0_ in 𝒲_*R*_ is chosen to be uniform with support on both positive and negative values. The Bayesian inference methodology of Section 2.5 is carried out with *M*^*\**^ = 50, 000 to obtain 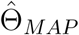 and samples 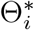 from the posterior of Θ in (19). The curves of *γ*_*o*_ *I*_*o*_(*t*) and Δ*R*_*o*_(*t*) *≡ R*_*o*_(*t*) *− R*_*o*_(*t −* 1) for each day *t* are obtained based on 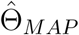 and are displayed in Figure 3. This submodel captures broad features (increasing and decreasing trends) of the reported cases trajectories in all three panels for Malaysia, Selangor and Sarawak. To quantify the overall change in transmission before and after MCO implmentation, Δ_*t*_ (see (15)) is obtained for *t* in 𝒲_*L*_ and 𝒲_*R*_. The plots of Δ_*t*_versus *t* are shown in Figure 4. A consistent feature of the plots in all three panels is that they first increase for time points *t ≤ T*^*\**^ followed by a significant drop for *t > T*^*\**^. Hence, we conclude that an exponential rise in cases occurred right after the completion of the Sri Petaling gathering on March 1st 2020, and the implementation of the MCO successfully stemmed the exponential rise at the national level, Selangor and Sarawak.

**Figure 3:**
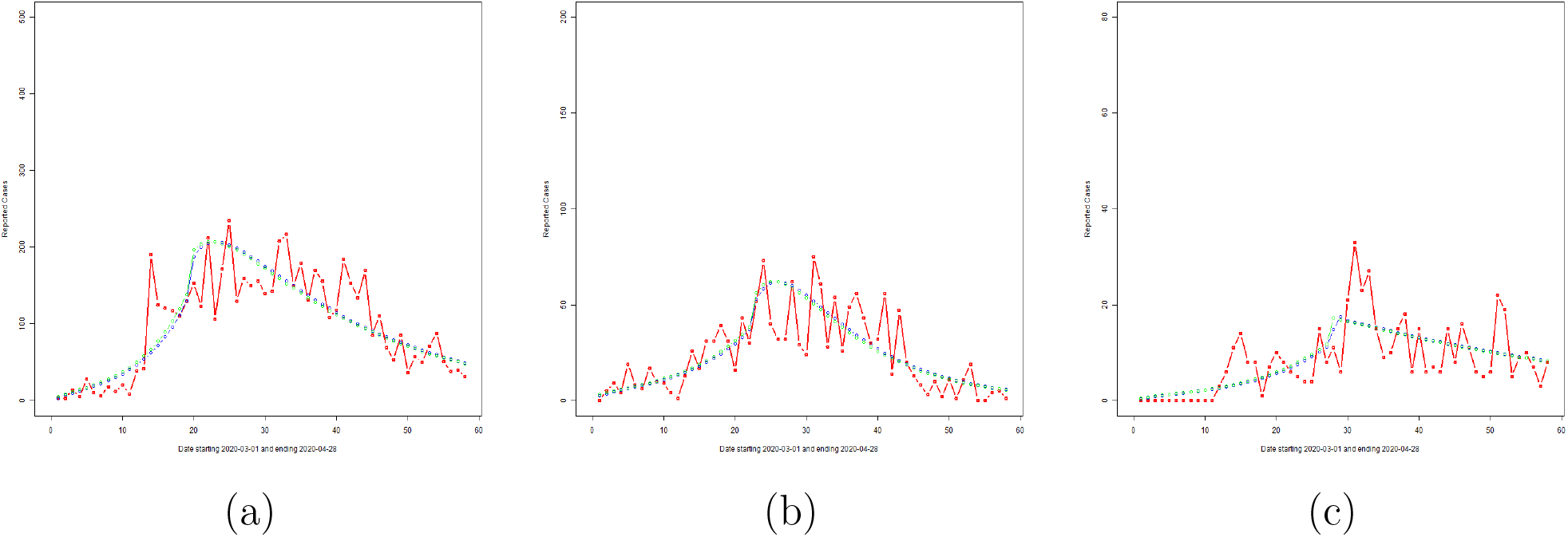
Reported daily cases (red line), and overlay plots of Δ*R*_*o*_(*t*) (blue line) and *γ*_*o*_ *I*_*o*_(*t*) (green line) based on 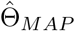 for the constant rate submodel for (a) Malaysia, (b) Selangor and (c) Sarawak.

**Figure 4:**
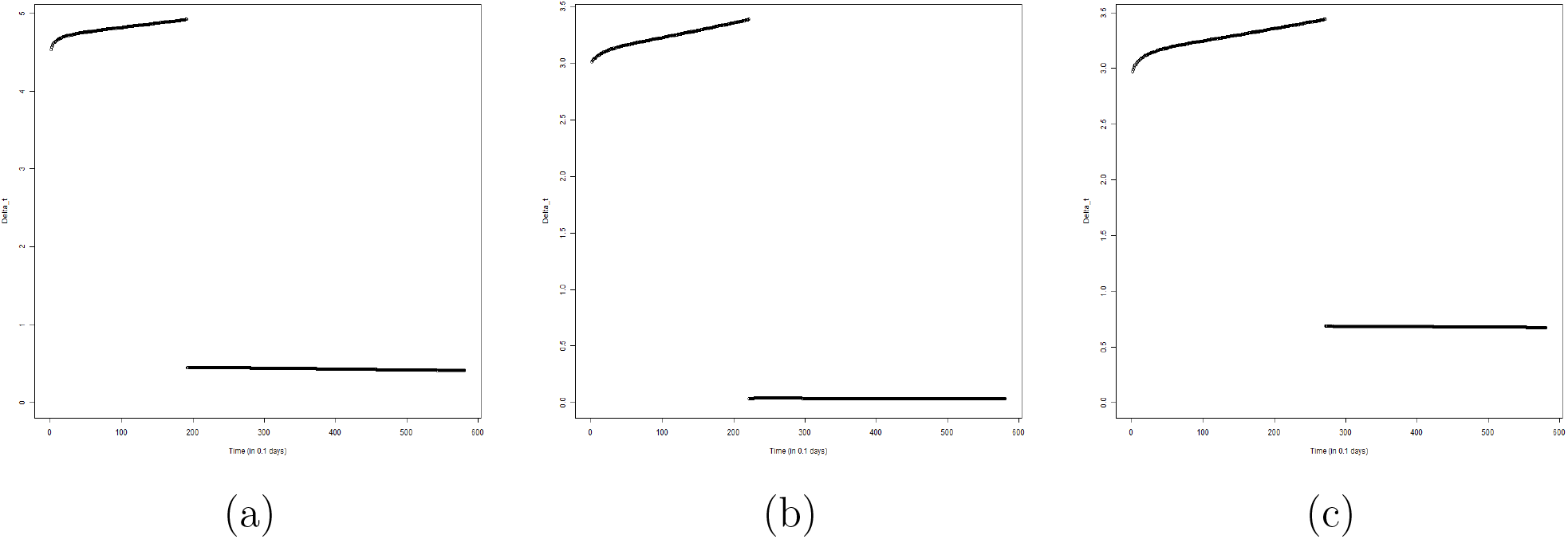
Plots of Δ_*t*_ versus *t* for the constant rate submodel for (a) Malaysia, (b) Selangor and (c) Sarawak.

With reduced disease transmission established in 𝒲_*R*_, we next proceed to utilize the functional form (13) of *β*_*o*_(*t*) as a quantitative model for transmission decay in 𝒲_*R*_. The Bayesian inference methodology of Section 2.5 is applied to the full model with *M*^*\**^ = 50, 000 to obtain 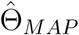 and samples 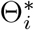 from the posterior of Θ in (19). The curves of *γ*_*o*_ *I*_*o*_(*t*) and Δ*R*_*o*_(*t*) *≡ R*_*o*_(*t*) *− R*_*o*_(*t −* 1) for each day *t* are obtained based on 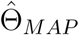 and are displayed in Figure 5. Daily cumulative cases and the curve of *R*_*o*_(*t*) are displayed in Figure 6. We note from these figures that the proposed model captures broad features of the observed data and is an improvement over the constant rate sub-model. Uncertainty estimates are obtained for all unknown parameters in Θ based on the ensemble 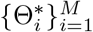. Variabilility estimates can be obtained for all parameters and their functions. As an illustration, we demonstrate the extent of variability inherent in the posterior visually for the expected *R*_*o*_(*t*) curve given by *γ*_*o*_ *I*_*o*_(*t*) (see (16)) for *t ∈* [*T*_0_, *T*_1_]. This is displayed in Figure 7 which shows that most of the reported case numbers are well within the limits of variability of the posterior. Hence, the proposed model together with the negative binomial likelihood are able to explain the variability in the reported case numbers. However, there are a few exceptions, the most notable being the reported case number on Day 14 for Malaysia in Figure 7(a). We present an explanation for this outlying case later in the Discussion section.

**Figure 5:**
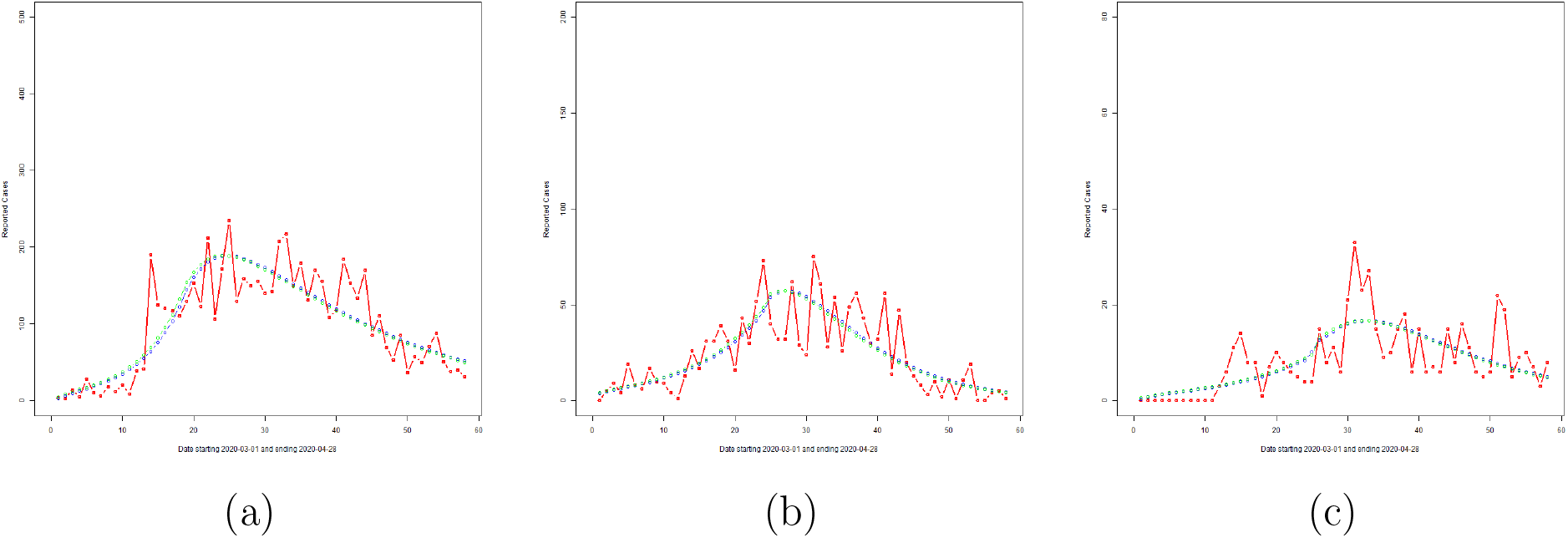
Reported daily cases (red line), and overlay plots of Δ*R*_*o*_(*t*) (blue line) and *γ*_*o*_ *I*_*o*_(*t*) (green line) based on 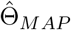 for (a) Malaysia, (b) Selangor and (c) Sarawak.

**Figure 6:**
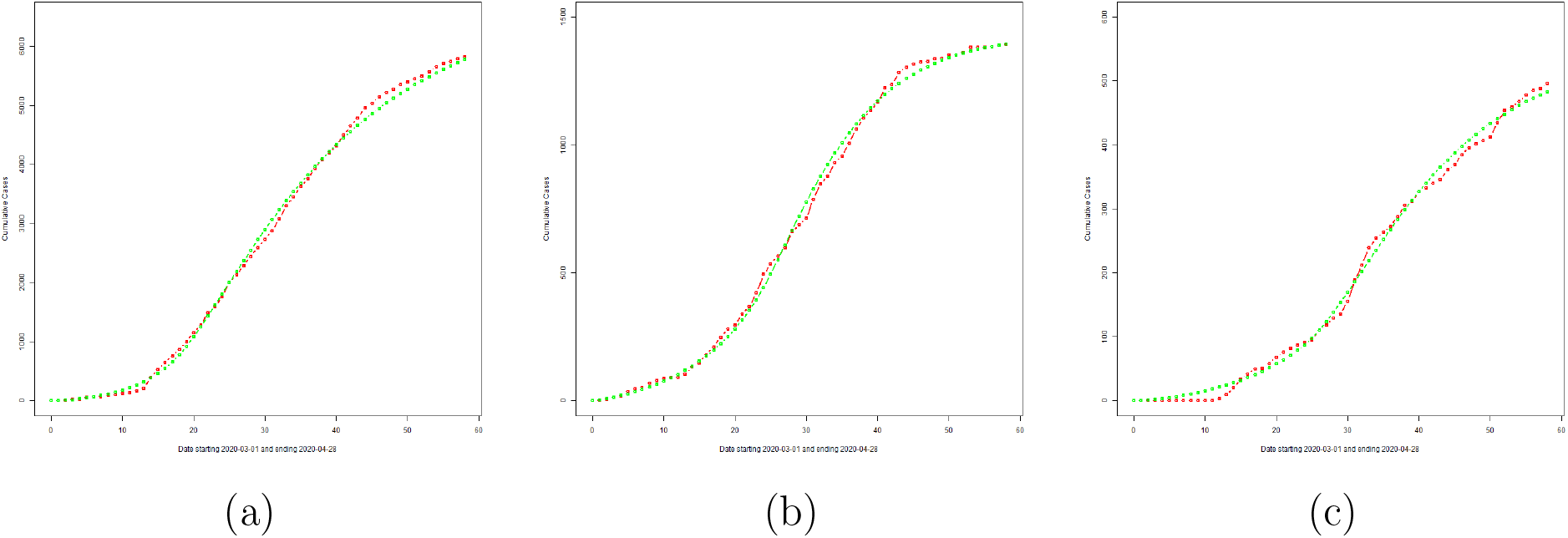
Reported cumulative cases (red line), and overlay plots of *R*_*o*_(*t*) (green line) based on the MAP estimate for (a) Malaysia, (b) Selangor and (c) Sarawak.

**Figure 7:**
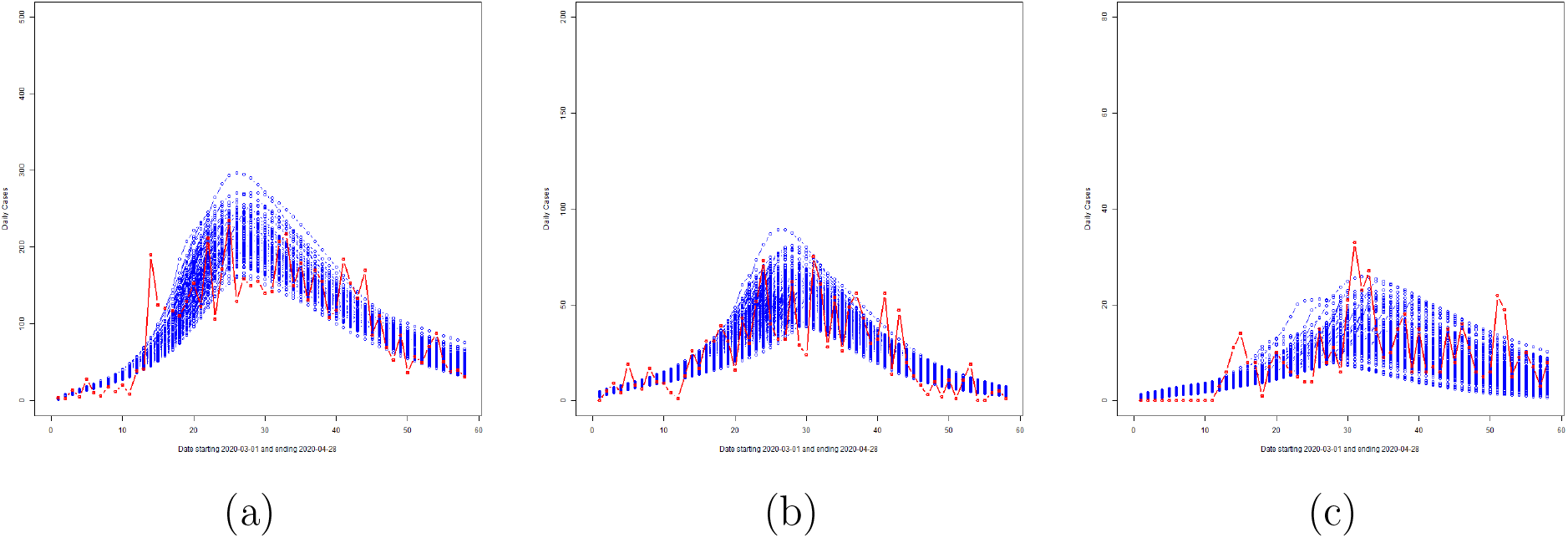
Illustration of the variability of *γ*_*o*_ *I*_*o*_(*t*) from the posterior: Reported cumulative cases (red line), and overlay plots of *γ*_*o*_ *I*_*o*_(*t*) for (a) Malaysia, (b) Selangor and (c) Sarawak.

Further results from the Bayesian analyses are summarized in Tables 1, 2 and 3. These tables give the MAP estimates of parameters and their corresponding 95% credible intervals for Malaysia, Selangor and Sarawak, respectively. We provide a summary of the salient findings here. The symptomatic and asymptomatic infectious periods as well as the incubation periods are found to be around 6-8 days for Malaysia, Selangor and Sarawak. These findings are similar to values reported in the literature for other countries; see, for example, [16, 17, 18, 27]. Change-points *T*^*\**^ are estimated not too far away from the date of MCO implementation, March 18th 2020. For Malaysia, *T*^*\**^ = 18 is the MAP estimate which corresponds to March 19th, 2020, and the associated 95% credible interval is (17, 21). For Selangor, the MAP estimate of *T*^*\**^ is *T*^*\**^ = 22 with (17.5, 25) being the 95% credible interval. For Sarawak, the transition date is less precise. The MAP estimate is *T*^*\**^ = 25 (March 26th 2020) but the 95% credible interval (18, 32.5) is much larger indicating higher uncertainty in *T*^*\**^. This can be attributed to the fact that the trajectory of reported case numbers for Sarawak shows a slower and more gradual increase, then decrease, compared to Malaysia and Selangor (see Figure 5).

**Table 1:**
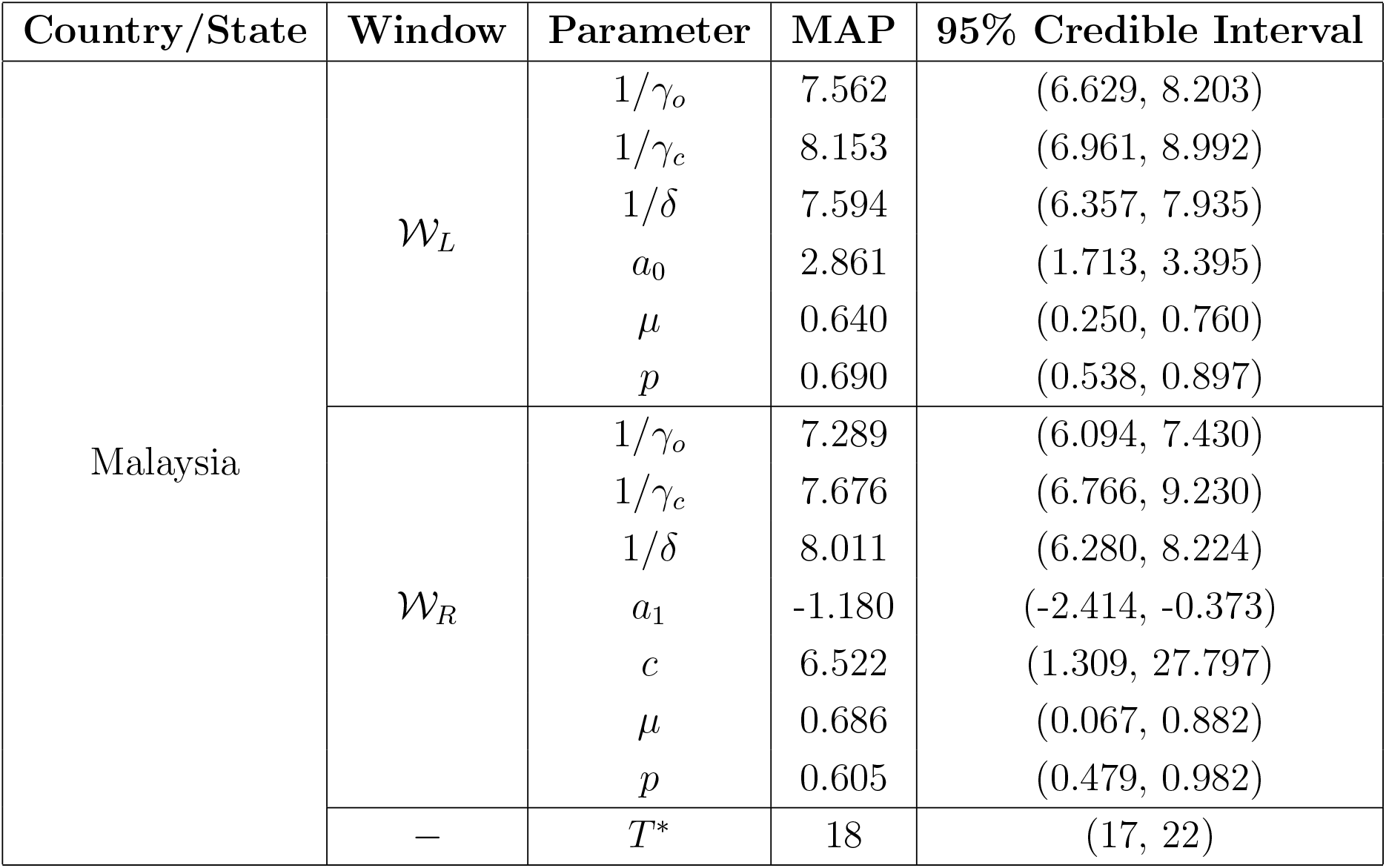
MAP estimates and associated credible intervals for Malaysia.

| Country/State | Window | Parameter | MAP | 95% Credible Interval |
| --- | --- | --- | --- | --- |
| Malaysia | $\mathcal{W}_L$ | $1/\gamma_o$ | 7.562 | (6.629, 8.203) |
| | | $1/\gamma_c$ | 8.153 | (6.961, 8.992) |
| | | $1/\delta$ | 7.594 | (6.357, 7.935) |
| | | $a_0$ | 2.861 | (1.713, 3.395) |
| | | $\mu$ | 0.640 | (0.250, 0.760) |
| | | $p$ | 0.690 | (0.538, 0.897) |
| | $\mathcal{W}_R$ | $1/\gamma_o$ | 7.289 | (6.094, 7.430) |
| | | $1/\gamma_c$ | 7.676 | (6.766, 9.230) |
| | | $1/\delta$ | 8.011 | (6.280, 8.224) |
| | | $a_1$ | -1.180 | (-2.414, -0.373) |
| | | $c$ | 6.522 | (1.309, 27.797) |
| | | $\mu$ | 0.686 | (0.067, 0.882) |
| | | $p$ | 0.605 | (0.479, 0.982) |
| | — | $T^*$ | 18 | (17, 22) |

**Table 2:** MAP estimates and associated credible intervals for Selangor.

| Country/State | Window | Parameter | MAP | 95% Credible Interval |
| --- | --- | --- | --- | --- |
| Selangor | $\mathcal{W}_L$ | $1/\gamma_o$ | 7.952 | (6.434, 8.320) |
| | | $1/\gamma_c$ | 8.611 | (6.826, 9.120) |
| | | $1/\delta$ | 6.747 | (5.615, 7.905) |
| | | $a_0$ | 2.041 | (1.296, 2.956) |
| | | $\mu$ | 0.468 | (0.211, 0.774) |
| | | $p$ | 0.785 | (0.519, 0.885) |
| | $\mathcal{W}_R$ | $1/\gamma_o$ | 7.342 | (6.100, 7.482) |
| | | $1/\gamma_c$ | 9.066 | (6.839, 9.099) |
| | | $1/\delta$ | 7.099 | (5.518, 8.033) |
| | | $a_1$ | -2.384 | (-2.953, -0.303) |
| | | $c$ | 4.037 | (0.144, 13.069) |
| | | $\mu$ | 0.374 | (0.177, 0.881) |
| | | $p$ | 0.708 | (0.486, 0.948) |
| | — | $T^*$ | 22 | (17.5, 25) |

**Table 3:** MAP estimates and associated credible intervals for Sarawak.

| Country/State | Window | Parameter | MAP | 95% Credible Interval |
| --- | --- | --- | --- | --- |
| Sarawak | $\mathcal{W}_L$ | $1/\gamma_o$ | 8.947 | (6.647, 9.567) |
| | | $1/\gamma_c$ | 11.399 | (8.936, 14.138) |
| | | $1/\delta$ | 7.251 | (7.056, 7.972) |
| | | $a_0$ | 1.627 | (1.139, 2.618) |
| | | $\mu$ | 0.250 | (0.027, 0.879) |
| | | $p$ | 0.735 | (0.502, 0.979) |
| | $\mathcal{W}_R$ | $1/\gamma_o$ | 7.158 | (6.195, 7.954) |
| | | $1/\gamma_c$ | 11.364 | (8.993, 14.278) |
| | | $1/\delta$ | 7.648 | (6.910, 8.287) |
| | | $a_1$ | -0.354 | (-1.896, -0.099) |
| | | $c$ | 0.540 | (0.168, 2.705) |
| | | $\mu$ | 0.433 | (0.013, 0.988) |
| | | $p$ | 0.880 | (0.521, 0.987) |
| | — | $T^*$ | 25 | (18, 32.5) |

The plots of Δ_*t*_ versus *t* for Malaysia, Selangor and Sarawak are provided in Figure 8. We note that all three panels in Figure 8 indicate a decay in the transmission rates after *T*^*\**^. The measure Δ_*t*_ is calculated to be approximately 3.55, 2.79 and 2.65 for Malaysia, Selangor and Sarawak, respectively, at the start of *𝒲*_*L*_, that is, when *t* = *T*_0_. Hence, Selangor achieved a rate of increase that is closer to the national level compared to Sarawak. Sarawak’s cases increased but at a much slower rate compared to Selangor and Malaysia. Starting from *t* = *T*_0_, Δ_*t*_ showed an increase in *𝒲*_*L*_, reaching values of 5.58, 3.35 and 2.96 at *t* = *T*^*\**^, respectively, for Malaysia, Selangor and Sarawak. After *T*^*\**^, Δ_*t*_ for Malaysia, Selangor and Sarawak registered a decay demonstrating the effectiveness of the MCO. Δ_*t*_ declined sharply to a value around 0.55, 0.01 and 0.37, respectively, for Malaysia, Selangor and Sarawak at *t* = *T*^*\**^ + 10, and after that, it declined more gradually to its corresponding asymptote. Based on Δ_*t*_, it is seen that the initial transmission rates tend to be higher for areas with a higher population density (comparing Selangor and Sarawak). On the other hand, based on the MAP estimates of *a*_1_ in 𝒲_*R*_ of *−*2.38 and *−*0.35 for Selangor and Sarawak, respectively (see Tables 2 and 3), higher population density areas also experience a faster decline in the transmission rates under an effective implementation of the MCO. Although the MAP estimate of *a*_1_ for Malaysia (*a*_1_ = *−*1.18 from Table 1) is not as negative as it should be, we will show in the next section that a redistribution of cases further improves this estimate of *a*_1_ and brings it closer to that of Selangor (see Section 4 for the details).

**Figure 8:**
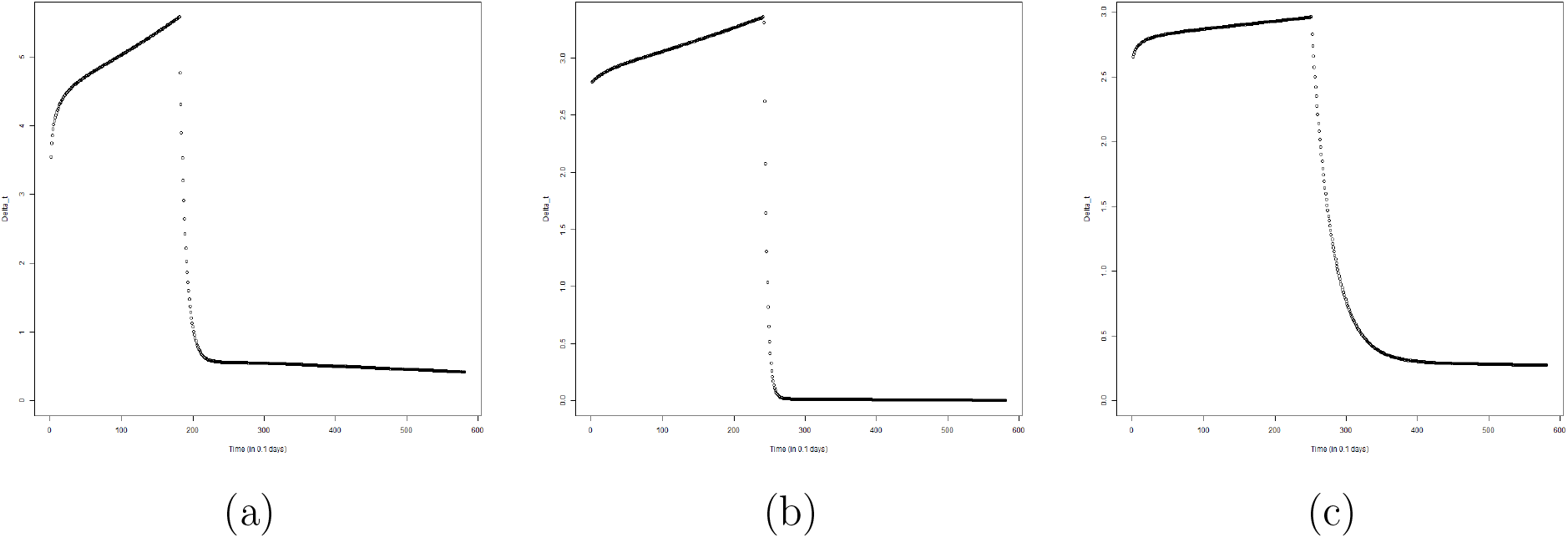
Plots of Δ_*t*_ versus *t*: Malaysia (a), Selangor (b) and Sarawak (c).

## 4 Discussion on Reporting Delays, Case Redistribution and Overdispersed Likelihoods

Figure 7 indicates the presence of outliers that fall outside the limits of variability of the posterior. The most notable outlier is the total number of new cases reported on Day 14 for Malaysia. Generally speaking, such outliers highlight a mismatch between the proposed model and the observed data, and point towards model inadequacy. However, we wish to emphasize that this is not the case here. One key consideration is the effect of delay, that is, whether or not the reported case numbers coincide with the day of testing. It is highly likely that a lag occurred in the reporting of cases since the COVID-19 experience was new to Malaysia. Based on the report [29], it is reasonable to assume that delays in testing and reporting were expected during the initial days of the COVID-19 outbreak in Malaysia. The peak on Day 14 seem to suggest a significant backlog of reporting of cases. The effects of reporting delays on observed case trajectories and parameter inference are illustrated here based on a simulation study. A delay-in-reporting model based on the multinomial distribution is assumed: Let *X ∼ Mult*(*D*_*t*_; *p*_1_, *p*_2_, *…, p*_*K*_), where *X* = (*X*_1_, *X*_2_, *…, X*_*K*_) with *K* = 5 and *X*_*k*_ is the number of cases (out of the total reported cases on day *t, D*_*t*_) that is to be redistributed to day *t − k* + 1 for *k* = 1, 2, *…, K*. The probabilities *p*_*k*_, *k* = 1, 2, *…, K* are chosen according to a truncated geometric distribution taking values in *k −* 1 for *k* = 1, 2, *…, K* with success probability 0.4. The cases redistribution model is applied to new cases reported from Day 10 until Day 15.The redistributed reported case trajectory, the best fit curves and associated variabilities are shown in Figure 9. Comparing Figures 7(a) and 9(b), one can immediately notice that the reported case numbers in Figure 9(b) are better explained by the variabilities of the underlying model and the negative binomial likelihood. Parameter estimates and credible intervals for the redistributed case numbers are given in Table 4. We present the salient findings here. Comparing Tables 1 and 4, we find that estimates of the infectious (both symptomatic and asymptomatic) periods have now become shorter. This is expected and reasonable since the model and likelihood do not have to account for the sudden steep rise in cases on Day 14 by preferring a larger infectious period. Nevertheless, the new infectious periods are still within the 6-8 day range and are consistent with previously reported literature. The redistribution of case numbers have also reduced the uncertainty around the MAP value of *T*^*\**^ = 18: The credible interval for *T*^*\**^ in Table 4 is narrower compared to that in Table 1. The MAP estimate of *a*_1_ is now *−*2.075, which is closer to that of Selangor compared to Sarawak.

**Table 4:** Summary results for Malaysia (with redistributed cases).

| Country/State | Window | Parameter | MAP | 95% Credible Interval |
| --- | --- | --- | --- | --- |
| Malaysia | $\mathcal{W}_L$ | $1/\gamma_o$ | 7.121 | (6.330, 8.223) |
| | | $1/\gamma_c$ | 7.694 | (6.829, 8.993) |
| | | $1/\delta$ | 7.379 | (6.211, 7.954) |
| | | $a_0$ | 3.018 | (1.643, 3.451) |
| | | $\mu$ | 0.709 | (0.218, 0.778) |
| | | $p$ | 0.810 | (0.545, 0.892) |
| | $\mathcal{W}_R$ | $1/\gamma_o$ | 6.821 | (6.058, 7.452) |
| | | $1/\gamma_c$ | 7.322 | (6.652, 8.921) |
| | | $1/\delta$ | 7.074 | (5.959, 8.195) |
| | | $a_1$ | -2.075 | (-2.888, -0.476) |
| | | $c$ | 8.438 | (0.675, 29.336) |
| | | $\mu$ | 0.621 | (0.105, 0.807) |
| | | $p$ | 0.756 | (0.540, 0.957) |
| | — | $T^*$ | 18 | (17, 21) |

**Figure 9:**
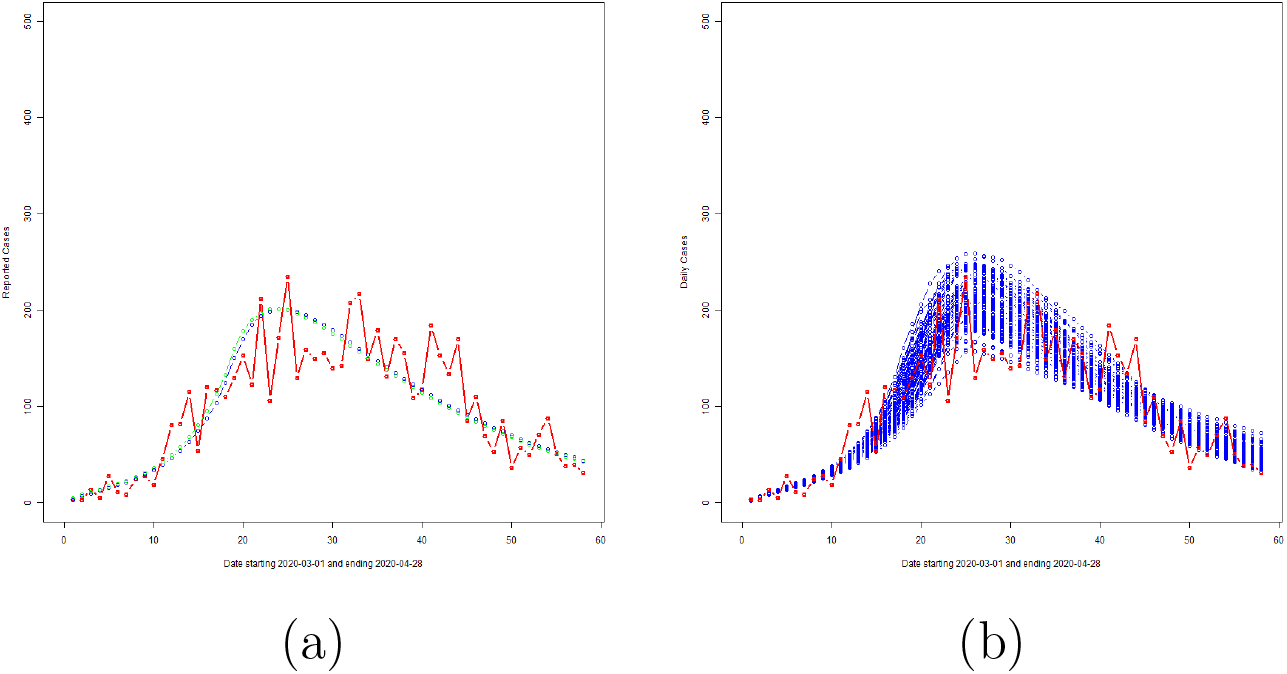
Effect of redistribution: Panel (a) shows the redistributed daily cases and the corresponding best fit curves of *γ*_*o*_ *I*_*o*_(*t*) (blue line) and Δ*R*_*o*_(*t*) (green line) based on 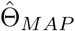. Panel (b) shows the variability of the fit based on the ensemble set 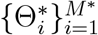.

A final point to be discussed is our preference for the negative binomial likelihood compared to the more traditional Poisson likelihood for modelling COVID-19 case numbers. Our initial investigation used the Poisson likelihood for reported case numbers but we found that the underlying model together with the Poisson likelihood was not able to capture inherent variabilities in the observed data. Hence, we opted for the overdispersed negative binomial likelihood which was able to satisfactorily represent the observed data via its overdispersion parameter *τ*. This is evidenced by the variability bands presented in Figures 7 and 9(b) which successfully enclose most of the reported case numbers. This coverage is further improved in Figure 9(b) by a redistribution of delayed cases. We also provide the loglikelihood values corresponding to the Poisson and negative binomial observation models in Table 5 for Malaysia (with original case numbers), Malaysia (with redistributed case numbers), Selangor and Sarawak. Note that the negative binomial log-likelihood values are consistently larger than the Poisson counterparts indicating a better model fit to observed data.

**Table 5:**
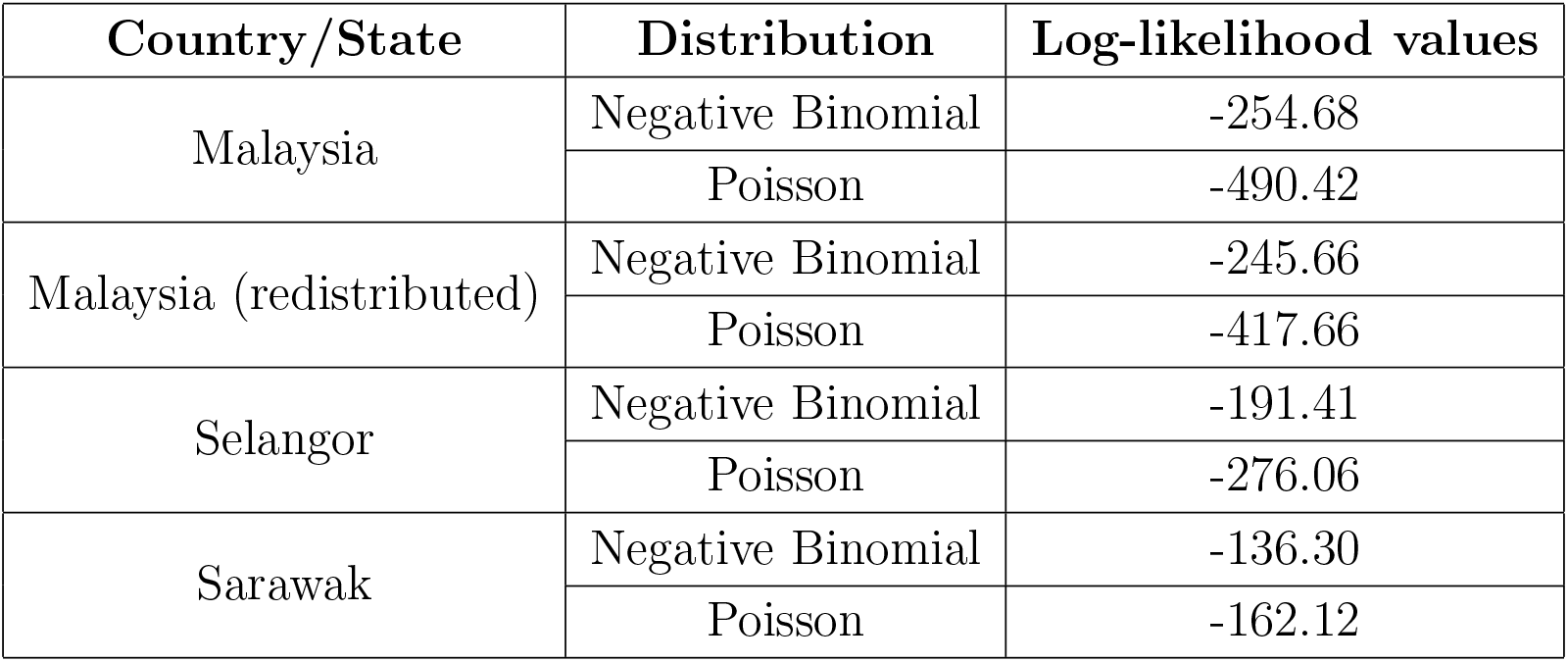
Loglikelhood values of the NB and Poisson likelihoods.

| Country/State | Distribution | Log-likelihood values |
| --- | --- | --- |
| Malaysia | Negative Binomial | -254.68 |
|  | Poisson | -490.42 |
| Malaysia (redistributed) | Negative Binomial | -245.66 |
|  | Poisson | -417.66 |
| Selangor | Negative Binomial | -191.41 |
|  | Poisson | -276.06 |
| Sarawak | Negative Binomial | -136.30 |
|  | Poisson | -162.12 |

## 5 Conclusion

Quantitative models and assessment of the impacts of the Sri Petaling gathering and implementation of MCO on COVID-19 spread in Malaysia are developed in this paper. The MCO implementation is found to be highly effective in containing (an exponential rise of) the COVID-19 outbreak in Malaysia. The analysis here quantitatively demonstrates how quickly transmission rates fall under effective NPI implemention within a short time period. Higher disease transmission is found in Selangor (a state with higher population density) compared to Sarawak. We also found that under MCO, the decline in transmission was faster in Selangor compared to Sarawak. The rise and fall of disease transmission in Selangor mirrored the national level whereas Sarawak showed a more gradual increase and decrease in COVID-19 transmission. The change points were mostly found to be close to the date of MCO implementation (18th March 2020) although Sarawak exhibited a larger uncertainty around that date due to its gradual and slower increasing and decreasing trends of reported case numbers. Our study developed a new model to represent COVID-19 spread in Malaysia that accounts for heterogeneity and asymptomatic transmissions. We found that reported case numbers in Malaysia exhibited large variabilities which can possibly be attributed to a delay in reporting, particularly during the early stages of the pandemic as the experience with handling COVID-19 was new to the country. Nevertheless, the model developed here together with the overdispersed negative binomial likelihood are able to capture salient features of COVID-19 spread in Malaysia and provide reliable quantitative assessments even under the challenges of limited and delayed data.

## Data Availability

The data is open source and can be accessed from these links: https://www.outbreak.my/ and http://covid-19.moh.gov.my/.

http://covid-19.moh.gov.my/

## Acknowledgment

The authors would like to acknowledge the Global Challenges Research Fund (GCRF) “Post MCO Strategies for Controlling Covid-19 in Malaysia” by the Scottish Funding Council and Heriot-Watt University. The authors declare no conflicts of interest. The authors would also like to thank the Director General of Health Malaysia for his permission to publish this article.

## Appendix: Prior Elicitation on Remaining Parameters

We describe the prior elicitation for the initial number of infectious and exposed individuals, *i*_0_ and *e*_0_, respectively, relevant only for 𝒲_*L*_. Conditional on *δ* and *γ*_*o*_, *i*_0_ (corresponding to observed, not cryptic) is given the prior elicitation 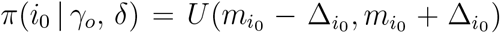 for some 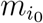 and 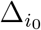. This prior, *π*(*i*_0_ | *γ*_*o*_, *δ*), on *i*_0_ is motivated from based on the differential equation for the *R*_*o*_ compartment. Note that 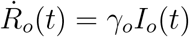 from (9), and hence, 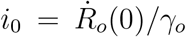. To obtain an estimate of 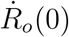, a second order polynomial is fitted using least squares to the trajectory of cumulative cases in a window of *m ≥* 3 days starting from 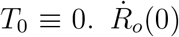 is then estimated by 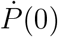 where 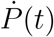 is the first derivative of the fitted polynomial, *P* (*t*). The mean of the uniform distribution on *i*_0_ is taken to be 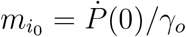. The initial number of exposed individuals, *e*_0_, is given the prior elicitation 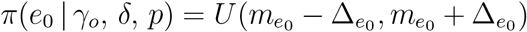 for some 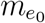 and 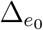. To find an expression for 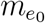, we rewrite (8) and note that *e*_0_ = (*İ*_*o*_(0) + *γ*_*o*_ *I*_*o*_(0))*/*(*p δ*). Next, substituting 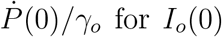 for *I*_*o*_(0), the mean of the uniform distribution on *e*_0_ is taken as 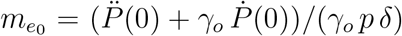 where 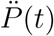 is the second derivative of *P* (*t*) with respect to *t*. The half-widths for both priors on *i*_0_ and *e*_0_ are taken as 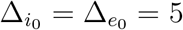. Thus, the initial prior distributions on *i*_0_ and *e*_0_ are based on the number of infectives and exposed in the original population; so they are un-normalized. This is because their estimates are calculated from reported case data. But these estimates are later normalized by the population size for input into the SIER and modified SEIR models.

The hyperparameters *a*_*ξ*_ and *b*_*ξ*_ for *ξ ∈ {µ, γ*_*o*_, *γ*_*c*_, *δ, p, w*_*i,o*_, 𝒲_*i,c*_, 𝒲_*s*_, *α}* are chosen based on values reported in previous studies where available. For example, the incubation period, defined as the period from being infected by COVID-19 to the onset of symptoms, is typically reported to be between 6 and 8 days on average [30]. Hence, we take *a*_*δ*_ = 6 and *b*_*δ*_ = 8 for the prior elicitation of 1*/δ*. For the infectious period, we consider 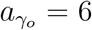 and 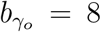 to encompass corresponding values available from the literature; see, for example, [16, 17, 18, 27]. The values of parameters reported in the literature are only taken as starting points.

